# Burnout in gastroenterology registrars: a pilot study of trainees in the East of England

**DOI:** 10.1101/2020.03.18.20038463

**Authors:** John Ong, Carla Swift, Wanyen Lim, Sharon Ong, Yasseen Al-Naeeb, Arun Shankar

## Abstract

**Objective:** The scale of burnout in UK gastroenterology trainees and the feasibility to determine its prevalence using the validated Maslach Burnout Inventory Human Services Survey (MBI-HSS) tool are unknown. A region-wide pilot study was conducted to determine the uptake of a 31-item questionnaire and estimate the prevalence of burnout in gastroenterology trainees within the East of England deanery (EoE). Symptom severity across the three domains of burnout (emotional exhaustion, depersonalisation, and low personal accomplishment), and frequently experienced stressors by gastroenterology trainees were also studied.

**Design:** This was a cross-sectional study involving gastroenterology trainees from 16 hospitals across EoE using a 31-item questionnaire. The questionnaire consisted of the 22-item MBI-HSS and 9 additional free-text questions. All gastroenterology trainees in EoE were invited to complete the anonymized survey online. Data were analysed quantitatively and qualitatively.

**Results:** Uptake of the survey was above-average; 44.0% (40/91) response rate. 57.5% (23/40) of gastroenterology trainees suffered emotional exhaustion. 23.5% (8/34) had depersonalisation and 63.9% (23/36) experienced low professional accomplishment. Burnout prevalence was 35.3% (12/34). Only 48.4% (15/31) of gastroenterology trainees were aware of professional support services within EoE. Stressors related to service requirements and professional relationships were commonly reported; 65.6% and 25.0% respectively.

**Conclusions:** It is feasible to use a 31-item questionnaire to detect and study burnout in a national cohort of gastroenterology trainees. Burnout in EoE gastroenterology trainees was high and this may reflect the national prevalence within the specialty. Larger studies, greater awareness of burnout, and better access to professional support services are needed.

**Summary Box:** *What is already known about the subject?:*

- Burnout in physicians is a growing problem worldwide which can lead to personal ill-health and suboptimal patient care.
- Burnout in young gastroenterology fellows in the US are reported as high as 50% but the prevalence in UK gastroenterology trainees is unknown.
- The Maslach Burnout Inventory - Human Services Survey (MBI-HSS) is the most validated tool to determine physician burnout but survey length may affect uptake by UK gastroenterology trainees and the feasibility of future studies.

*What are the new findings?:*

- This pilot study demonstrated the feasibility of a 31-item questionnaire which included the MBI-HSS in studying burnout in UK gastroenterology trainees.
- Emotional exhaustion and a sense of low personal accomplishment affect more than half of gastroenterology trainees within the East of England.
- The prevalence of burnout in UK gastroenterology trainees is estimated to be high (35.3%) but larger studies are needed.
- Approximately half of gastroenterology trainees in the East of England were not aware of existing support services to help them cope with burnout.

*How might it impact on clinical practice in the foreseeable future?:*

- This pilot study may increase the awareness of burnout among UK trainees and trainers in gastroenterology.
- An estimate of burnout prevalence in UK gastroenterology trainees is provided so future research and remediation measures in the specialty can be justified.

## INTRODUCTION

Burnout is an increasingly recognised occupational hazard among clinicians worldwide. It recently has been recognized as an “occupational phenomenon” in the 11th revision of the International Classification of Disease (ICD-11), and globally 30-50% of clinicians are estimated to have symptoms of burnout.[1] It is typically characterized by symptoms across three domains: emotional exhaustion, depersonalization and a sense of low personal accomplishment (self-worth); the latter two domains have recently been referred to as “cynicism” and “reduced professional efficacy” respectively.[2] These symptoms exist on a scale of varying severity and the lack of awareness of its nature can lead to its under-recognition.[3] Unaddressed, clinician burnout can have a negative impact on patient outcomes through impaired professionalism, poor communication, decreased patient satisfaction, professional errors, near misses and even patient harm.[1] To clinicians, burnout may also cause depression and suicidal ideations, sleep disturbances, alcoholism, musculoskeletal disorders, hypertension and ischaemic heart disease.[4-7] Nonetheless, burnout is reversible and well placed support mechanisms are vital in the timely intervention and preservation of mental well being of clinicians in distress, especially those in the early stages of their careers.

Gastroenterology is a busy speciality often involving high workloads and large patient volumes, especially in the public sector. Unsurprisingly, the prevalence of burnout in gastroenterology fellows and young gastroenterologists have been reported as high as 54% in the US.[3,8] In the UK, gastroenterology training is equally demanding; trainees often hold many responsibilities involving general medical on calls, patient care in 30-40 bedded speciality wards, inpatient consults, outpatient clinics, endoscopy lists, speciality licensing exams, and for some, research commitments. Importantly, the prevalence of burnout in UK gastroenterology trainees has not been previously determined and the scale of the problem remains unknown.

Several tools have been developed to detect burnout. These include, but are not limited to the 22-item Maslach Burnout Inventory Human Services Survey (MBI-HSS), the 19-item Copenhagen Burnout Inventory, and the 16-item Oldenburg Burnout Inventory. The MBI-HSS is by far the most extensively validated and widely used tool in medical professionals.[9,10] However, when the 22-item MBI-HSS is supplemented with additional questions (e.g. demographic data collection) as is often the case, resulting questionnaires are usually extensive and yield low response rates.[11] We designed a 31-item questionnaire which contained both the 22-item MBI-HSS and 9 free-text questions. It was unknown if the response rate from UK gastroenterology trainees to this long questionnaire would be adequate for meaningful data analysis; typical response rates for US-based studies of gastroenterology fellows and gastroenterologists range from 8.1% to 12%.[8,12,13] Furthermore, a recent 64-item survey on stress in UK gastroenterologists yielded a response rate of only 29%.[14]

We hypothesized that burnout could be accurately detected using the MBI-HSS and survey response rates could be optimised by using streamlined questions. The objectives of this pilot study were to: (i) evaluate the uptake of a burnout questionnaire by response rates in a local trainee cohort before extending this study nationally, (ii) obtain an estimate of burnout prevalence within the East of England deanery (EoE), and (iii) understand common stressors that UK gastroenterology trainees face. A response rate of >30% was defined as acceptable since this is higher than typical response rates reported in un-incentivised studies of burnout.[15]

For the benefit of readers not familiar with the UK gastroenterology training system, we use the terms “gastroenterology registrars” and “gastroenterology trainees” interchangeably for better clarity. Standard specialty training in gastroenterology within the UK spans 5 years of fulltime clinical training (ST3, ST4, ST5, ST6 and ST7).

## METHODS

### Design and administration of the questionnaire

A two-part questionnaire was designed to detect the presence of burnout and collect qualitative data for further analysis. The first part comprised of the 22-question MBI-HSS which assessed burnout symptoms in the three domains of emotional exhaustion, depersonalisation (or cynicism) and a sense of low personal achievements (or professional efficacy). The second part comprised of 9 free-text questions.

For the 22 questions that related to the MBI-HSS, each question was graded on a seven-point Likert scale according to the frequency of symptoms, ranging from “never” (0) to “every day” (6). The Cronbach alpha in each domain was greater than 0.7. Scores from questions within each domain were summated and the average score was calculated for each respondent. Respondent scores were assessed for abnormal values which were provided in the proprietary MBI Manual 4th Edition (Method 2).[16] Abnormal values (proprietary) were derived from a population of 6,269 healthcare workers using the following formulae: Abnormal EE = Mean + (SD*0.5), Abnormal DP = Mean + (SD*1.25), and Abnormal LPA = Mean + (SD*0.1).[16] The higher the summated score or average score in the emotional exhaustion (EE) and depersonalisation (DP) subscales, the more frequent and severe the symptoms. Conversely, in the subscale measuring low personal accomplishment (LPA), the lower the summated score or average score, the more severe the symptoms. Individual summated scores were used to determine burnout (Method 1).[16] Clinical burnout was defined using the Maslach recommended criteria as the presence of either a high summated EE score with a high summated DP score, or a high summated EE score with a low summated LPA score (summated EE ≥ 27 and summated DP ≥ 13, or summated EE ≥ 27 and summated LPA ≤ 31).[17]

The 9 free-text questions (Supplementary Table 1) in the second part of the questionnaire gathered demographic data as well as information on the most significant stressors that trainees perceived. One question on General Internal Medical (GIM) on calls was included because it is widely accepted that the acute medical registrar role is a highly demanding and stressful responsibility within the UK [18], and therefore may have had an effect on burnout rates.

A license to reproduce the MBI-HSS was obtained (www.mindgarden.com) and the MBI-HSS questions, together with the supplementary questions, were transcribed to an online platform (www.surveymonkey.co.uk). For the ease of processing raw data, the entire questionnaire was divided into four pages; page 1 contained 9 questions on EE, page 2 contained 8 questions on LPA, page 3 contained 5 questions on DP, and page 4 contained the 9 free-text questions. An electronic link to the questionnaire was then circulated to all 91 gastroenterology trainees within EoE via work and personal email, and data was collected between 15 January 2020 and 15 February 2020. A reminder email was sent on 29 Jan 2020. All responses were anonymised. Phone reminders were not utilised to preserve respondent anonymity.

Data from the MBI-HSS and parts of supplementary questions were analysed quantitatively. The remaining questions in the second part of the questionnaire were analysed qualitatively. Trainees were asked for the most significant job-related factor that contributed to their stress levels, and their responses were grouped under the most appropriate of the four themes: service requirements (workload, staffing levels etc), professional relationships (with colleagues and patients), training (e.g. exams, length of training, program requirements) and others.

### Statistical analyses

MedCalc 19.1.5 was used to perform the statistical analyses. Trainee variables were tested for normality using the Shapiro-Wilk method where applicable.[19] Parametric results were reported as mean ± standard deviation and non-parametric results were reported as median and inter-quartile range (IQR). Age of trainees between burnt out and non-burnt out groups were compared using the Mann-Whitney U test. When numbers were greater than five in a contingency table, the chi-squared test was used to test for differences in burnout proportions between groups; training grades and training centres (tertiary centre, district general hospital and research centre). When trainee numbers were small (n ≤ 5) in a contingency table, Fisher’s exact test was used to calculate p-values for better accuracy; employment status (full-time vs. less than full-time), gender (male vs. female), and GIM on calls (yes vs. no).

Two-tailed *p* values were reported for all tests and the significance level was set at 95%. Bonferroni correction was applied for multiple hypothesis testing if a significant p-value was obtained. Regression models were not used because the small sample size would not provide accurate results for meaningful interpretation.[20] A logistic regression model to identify relationships between trainee associated variables and burnout will be used in a planned national study where sample size would be considerably larger.

### Ethics

Prior to designing the survey, the authors completed the Medical Research Council and NHS Health Research Authority decision tool (www.hra-decisiontools.org.uk) which determined ethical approval from a local research ethics committee (REC) was not required (Appendix 1). All participants were automatically anonymised by the online survey platform and trainees were made aware of this in their invitation email. Trainees were also informed the survey was for research purposes and participation was voluntary. Completion of the survey conferred implied consent and the authors only received anonymised responses with no trainee identifiable information. There were no risks posed to participants and participation in the survey was not incentivised.

### Patient and Public Involvement

This study did not involve any members of the public or patients.

### Exclusion Criteria And Missing Data

Missing data from incomplete DP and LPA questions were not used in the calculation of burnout rates. Average DP and LPA scores were calculated from incomplete DP and LPA questions, as permissible and advised by product literature. Missing data from the second part of the survey were not included in the respective fields of analysis. All authors in this paper were excluded from this study.

## RESULTS

### Response rates

The uptake of the questionnaire was good, achieving a response rate of 44.0% (41/90). 62.5% (25/40) of all responses were received within the first 2 weeks of the survey, 35% (14/40) of responses were received within 1 week of the reminder email being sent and 2.5% (1/40) responded in the last week of the survey. There was a 100% (40/40) completion rate for questions on EE, 90.0% (36/40) completion rate for questions on LPA, and 85.0% (34/40) completion rate for questions on DP. For the second part of the questionnaire, 65.0% (26/40) of respondents completed all 9 questions.

### Demographics of respondents

Most gastroenterology trainees that responded to the survey were aged between 30-35 years old with the mode of respondents being ST6 (i.e. the fourth year out of five years of standard gastroenterology training). There were more male than female respondents however this is reflective of the number of female gastroenterologists including gastroenterology trainees within the UK (21-39%)[21]. There were slightly more gastroenterology trainees who were working in district general hospitals (DGHs) than tertiary centres: 43.8% vs. 53.1%. Most of the gastroenterology trainees were involved in GIM on calls. A summary of the demographic data is displayed in Table 1.

**Table 1:**
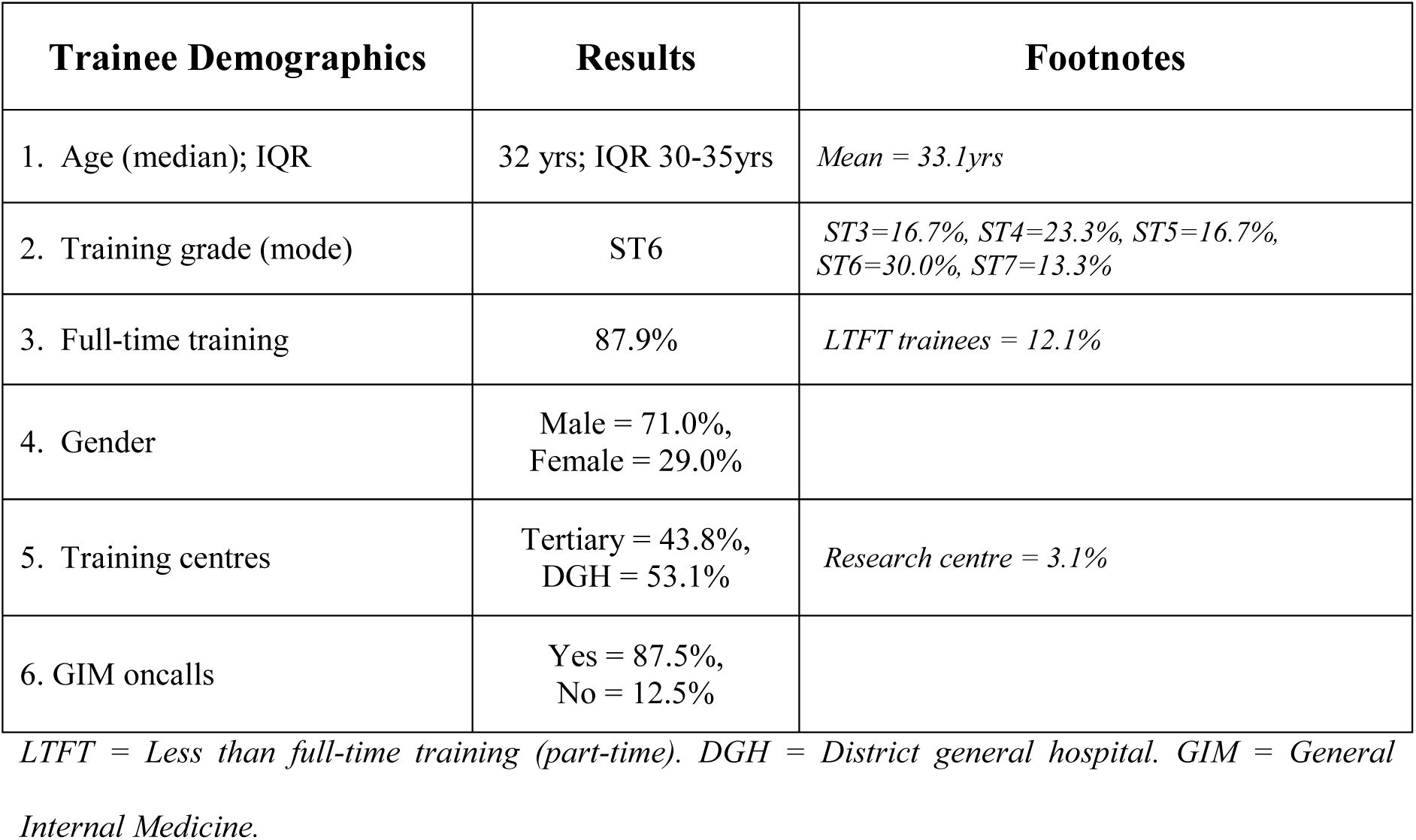
Demographic data of EoE gastroenterology trainees who responded in the survey.

| **Trainee Demographics** | **Results** | **Footnotes** |
| --- | --- | --- |
| 1. Age (median); IQR | 32 yrs; IQR 30-35yrs | *Mean = 33.1yrs* |
| 2. Training grade (mode) | ST6 | *ST3=16.7%, ST4=23.3%, ST5=16.7%, ST6=30.0%, ST7=13.3%* |
| 3. Full-time training | 87.9% | *LTFT trainees = 12.1%* |
| 4. Gender | Male = 71.0%,<br>Female = 29.0% |  |
| 5. Training centres | Tertiary = 43.8%,<br>DGH = 53.1% | *Research centre = 3.1%* |
| 6. GIM oncalls | Yes = 87.5%,<br>No = 12.5% |  |

*LTFT = Less than full-time training (part-time). DGH = District general hospital. GIM = General Internal Medicine.*

### The estimated prevalence of burnout and burnout related symptoms were high in gastroenterology trainees

Symptoms of burnout were high in gastroenterology trainees (Figure 1). Emotional exhaustion was present in 57.5% (23/40) of all respondents (EE mean 3.9. ± 0.8). Of these, 43.5% (10/23) experienced symptoms once a week or more frequently. Depersonalisation was detected in 23.5% (8/34) of all trainees (DP mean 3.9 ± 0.6), of these, 62.5% (5/8) experienced symptoms once a week or more frequently. 63.9% (23/36) of all respondents experienced a sense of low personal accomplishment (LPA mean 3.8 ± 0.7), of these, 52.2% (12/23) felt some sort of job satisfaction or fulfilment once a week or less. Overall, burnout was present in 35.3% (12/34) of gastroenterology trainees. Despite the high prevalence of burnout symptoms, only 48.4% (15/31) of gastroenterology trainees were aware that there were professional support services within the deanery to help trainees in distress.

**Figure 1.**
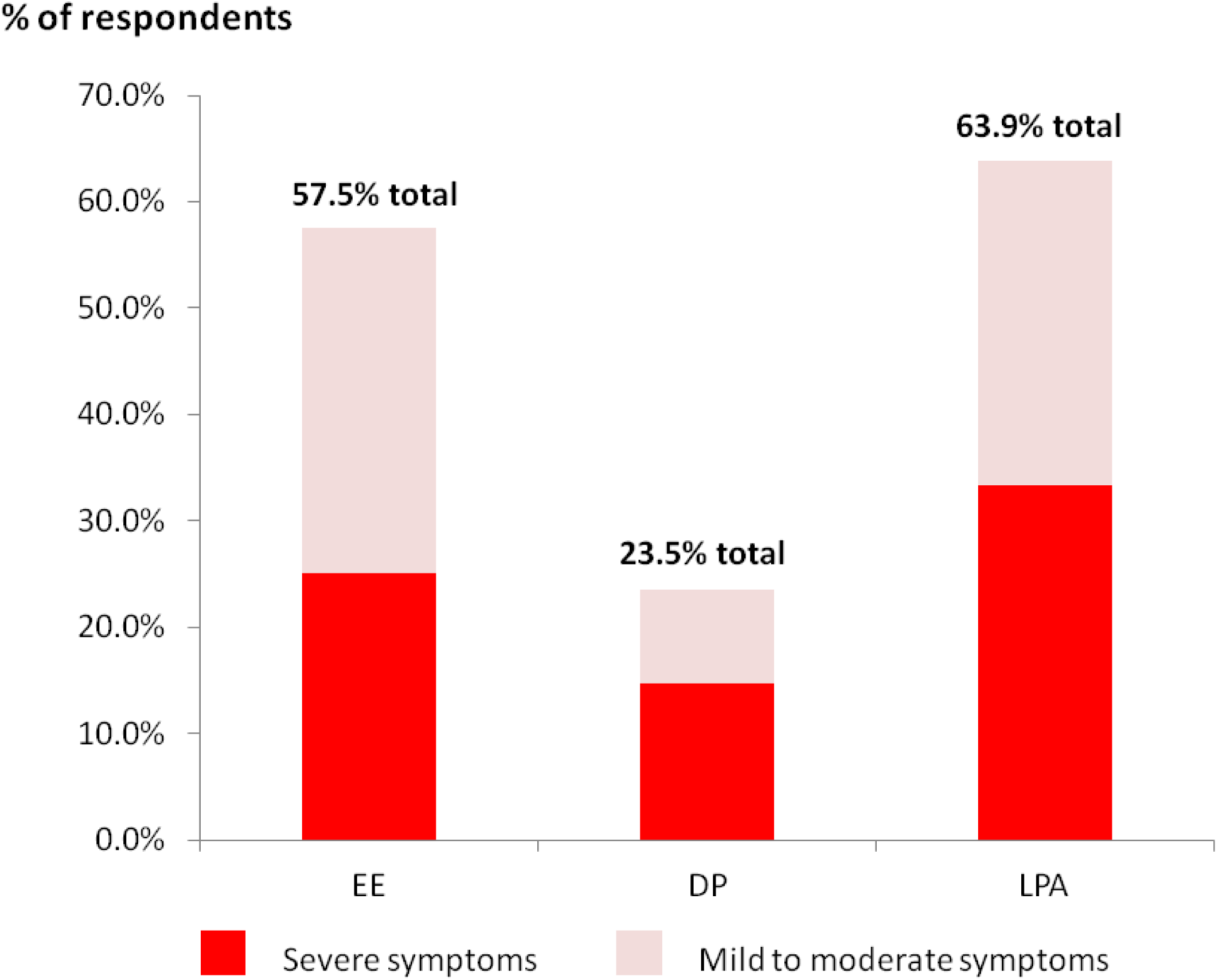
Frequency of burnout symptoms among gastroenterology trainees in the EoE

### Characteristics of burnt-out gastroenterology trainees

A summary of the characteristics between burnt out and non-burnt out gastroenterology trainees is displayed in Table 2. Briefly, burnt out gastroenterology trainees were observed to be slightly older, in later stages of training (ST5 and ST6 training grades), in full-time employment, working in tertiary centres, and did GIM on calls, however, none of these differences had any statistical significance when tested. For research purposes, the summated sub-scale scores were calculated and the results for the burnt-out group were as follow: EE=34.0 (median: 32, IQR 29-40), DP=16.3 ± 7.4, and LPA=28.2 ± 7.2. For the non-burnt group, the average of summated sub-scale scores were EE=21.2 ± 10.8, DP=7.6 (median: 6, IQR 5-10), and LPA = 38.0 ± 5.4.

**Table 2:** Characteristics of burnt out and non-burnt out gastroenterology trainees.

| Trainee variable | Burnt out<br>(a) | Non-burnt out<br>(b) | (a) vs. (b) |
| --- | --- | --- | --- |
| Age (mean) | 34.1 yrs | 32.6yrs | $p = 0.11$ |
| Grade<br>ST3<br>ST4<br>ST5<br>ST6<br>ST7 | 20.0% (1/5)<br>14.3% (1/7)<br>80.0% (4/5)<br>55.6% (5/9)<br>25.0% (1/4) | 80.0% (4/5)<br>85.7% (6/7)<br>20.0% (1/5)<br>44.4% (4/9)<br>75.0% (3/4) | $p = 0.12$ |
| Employment status<br>Full-time<br>LTFT | 41.4% (12/29)<br>0% (0) | 58.6% (17/29)<br>100% (4/4) | $p = 0.27$ |
| Gender<br>Male<br>Female | 40.9% (9/22)<br>33.3% (3/9) | 59.1 % (13/22)<br>66.7% (6/9) | $p = 0.70$ |
| Training Centre<br>Tertiary<br>DGH<br>Research centre | 42.9% (6/14)<br>29.4% (5/17)<br>100% (1/1) | 57.1% (8/14)<br>70.6% (12/17)<br>0% (0/0) | $p = 0.27$ |
| GIM on calls<br>Yes<br>No | 32.1% (9/28)<br>50.0% (2/4) | 67.9% (19/28)<br>50.0% (2/4) | $p = 0.49$ |

### Common stressors in UK gastroenterology trainees

87.1% (27/31) of gastroenterology trainees stated that job-related factors contributed to the most amount of stress in their lives, 12.9% (4/31) reported personal factors - family relationships and personal finances. Figure 2 illustrates the frequency of job-related stressors reported by gastroenterology trainees according to the themes. 65.6% (21/32) of trainees reported stressors related to service requirements, these comprised of workload (56.2%) and inadequate staffing levels (9.4%). Professional relationships were the second most frequently reported theme at 25.0% (8/32); these consisted of difficult and unsupportive colleagues (9.4%), expectations by seniors (9.4%), difficult patients (3.1%), and expectations of patients (3.1%). Stressors relating to training requirements were reported at a frequency of 12.5% (4/32) which comprised speciality exams (6.3%), length of training (3.1%) and curriculum requirements (3.1%). Other stressors that were reported include long commuting times between home and work (6.3%) and length of speciality training (3.1%).

**Figure 2:**
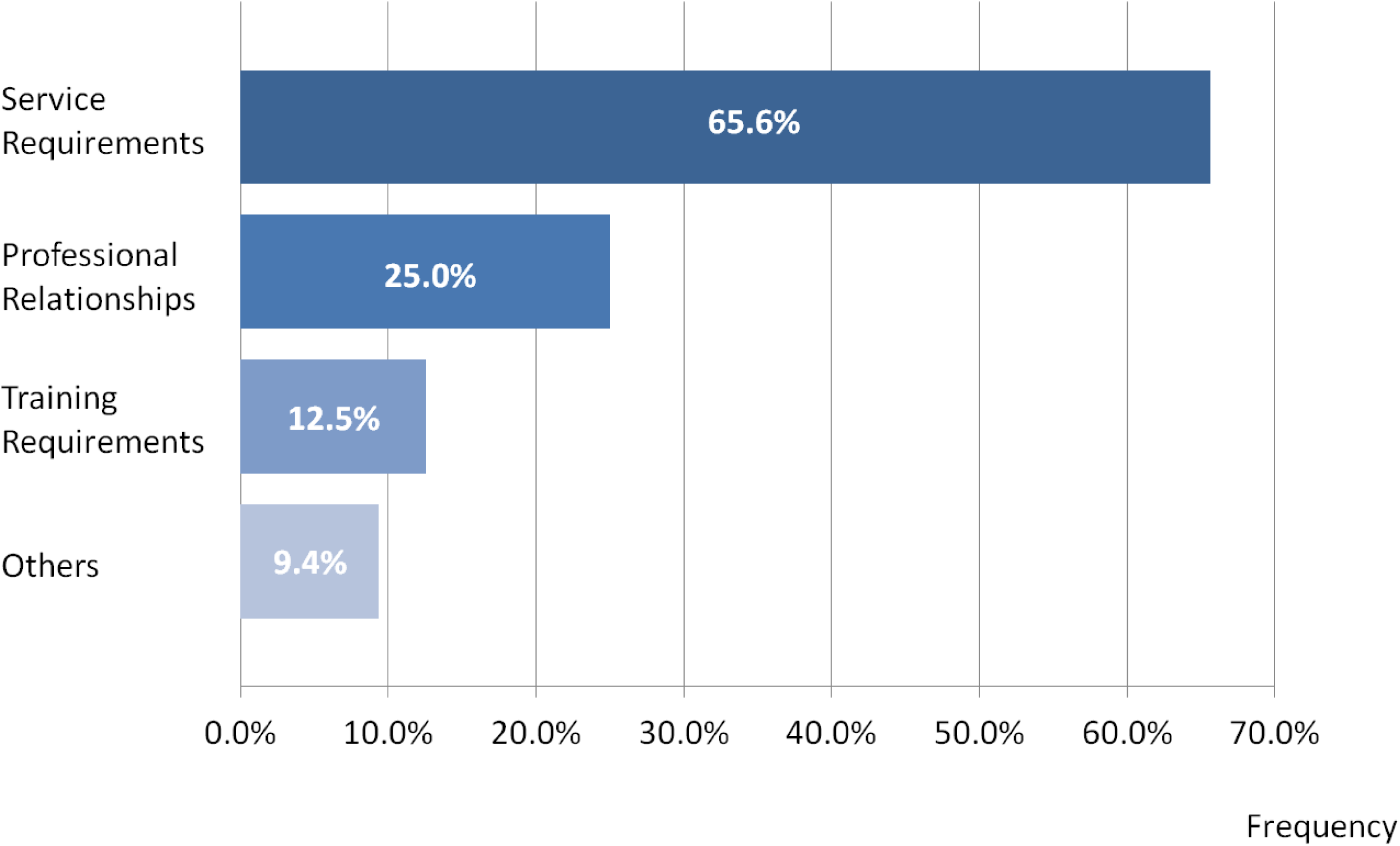
The frequency of significant stressors in gastroenterology trainees grouped by common themes.

## DISCUSSION

This study has used the 22-item MBI-HSS to detect burnout although abbreviated versions (aMBI) of the tool exist. These include the 2-item, 9-item MBI or 12-item aMBI which are favoured by some researchers because they are easier to administer and can yield better response rates. However, we have chosen not to use an abbreviated version of the MBI because these can be unreliable; we have previously demonstrated that the 12-question MBI had a poor positive predictive value of 33.3% (95% confidence interval:27.5-39.8%) and could overestimate the prevalence of burnout.[22] The study period was also capped at 1 month because given our experience in burnout studies of similar sample size, most of the responses were usually obtained within the first two weeks of the survey.[22] This is clearly evidenced in our observations of the current study as shown in the results. For the planned nationwide study, the study period will be extended to 3 months since the cohort size would be exponentially larger.

Even though the MBI-HSS is a well established tool, many researchers have adopted different criteria as well as numerical cut-off values to define burnout. Such heterogeneity in burnout definitions, as well as burnout tools, has precluded the accurate estimation of burnout in systematic reviews and meta-analyses.[9,23] In this study, burnout was defined by the Maslach-recommended criteria of “high EE and high DP” or “high EE and low PA” since this is the only definition that has been shown to have a clinical correlation; researchers have established that the combination of high scores in these domains correlate closely to work-related neurasthenia as defined by ICD-10.[16,17,24] Using this definition, the prevalence of clinical burnout was determined to be 35.3% but this is possibly an underestimation because burnt out trainees may not have engaged in this study.

Nonetheless, this study has demonstrated that the uptake of the 22-item MBI-HSS within gastroenterology trainees was good, achieving a response rate of 44.0% which is higher than commonly reported response rates in burnout studies. Interestingly, although a 100% completion rate was achieved in the emotional exhaustion questions (page 1 of 4 on the online survey), the completion rate gradually declined as the questionnaire progressed: 90% for LPA questions (page 2), 85.0% for DP questions (page 3) and 65.0% for free-text questions (page 4). In a planned nationwide study, the authors will consolidate all questions onto 2 pages to improve the completion rate; the first page to contain the 22-item MBI-HSS and the second page to contain free-text questions.

In our results, burnt out trainees were observed to be slightly older (34.1yrs vs. 32.6yrs), in ST5 and ST6 training grades, in fulltime employment, working in tertiary centres, and did GIM on calls. However, these differences did not have any statistical significance when tested. The lack of statistical significance, however, does not imply the lack of association but rather could be a result of the small sample size. In the planned nationwide study, these factors could be analysed more meaningfully in a larger sample size with regression methods to test for associations. Also, it was unexpected that difficult and unsupportive colleagues would be the second most frequently reported stressor (9.4%), jointly ranked with expectations by seniors (9.4%) and poor staffing levels (9.4%). The two former stressors may be addressed by interventions such as mentoring and mindfulness-based exercises [25-28], however the latter may prove to be more difficult for reasons discussed below.

Within the UK, the demand for gastroenterology services is gradually increasing but almost half of advertised gastroenterology consultant posts remain unfilled.[21] Several years are needed to train or recruit manpower to fill these vacancies and it is therefore unsurprising that current gastroenterology consultants and trainees face heavy workloads, and as a result, may experience high levels of stress and burnout. Recently, a survey on stress among UK gastroenterologists found that 48% considered leaving their current hospital of work, 44% had lost their temper at work and 6% had contemplated suicide.[14] These strongly suggest further studies in burnout are needed, especially in gastroenterology trainees and young consultants since early stage burnout can diminish the size of the future workforce, negatively impacting its sustainability, and have detrimental effects on the health of both patients and clinicians. More importantly, we have found that only 48.4% of all gastroenterology trainees were aware of existing professional support services within EoE deanery to help trainees in distress. As a result, bespoke lectures on well being and wider advertisement of support services have been planned for gastroenterology trainees within EoE since the completion of the study.

Nonetheless, the scale of burnout within gastroenterology in the UK remains unknown and must first be understood before any meaningful interventions can be designed. Once a nationwide study of gastroenterology trainees has been completed, the authors intend to conduct similar studies involving gastroenterology consultants, nurses and allied health professionals that constitute the rest of the UK gastroenterology workforce. In so doing, burnout prevalence amongst the workforce, awareness of, and access to support services across the country could also be studied.

This pilot study has several limitations. Being a voluntary survey, this study may have been affected by response bias and non-response bias although these are inherent problems of survey-based burnout studies. Also, the small sample size may not be fully representative of the gastroenterology trainees throughout the UK therefore a larger study is needed to accurately determine the prevalence of burnout. Although the MBI-HSS is the most reliable and validated tool to detect burnout to date, its limitation should be recognised because burnout syndrome itself is poorly characterized. Lastly, working hours and rota patterns in gastroenterology and GIM were not studied and therefore limited insight can be gained into the working environment of gastroenterology trainees. The authors acknowledge this was a compromise to achieve better response rates. Even though associations between longer working hours and burnout are known, the relationship is non-linear, often complex, and is influenced by many other factors such as personality traits (resilience) which are difficult to measure.

## Conclusion

It is feasible to use a 31-item questionnaire comprising the MBI-HSS to determine the prevalence of burnout in gastroenterology trainees in a larger cohort study. Most gastroenterology trainees within EoE have symptoms of burnout and the prevalence of burnout was estimated at 35.3%, however, larger scale studies are needed. Service requirements and professional relationships contribute the most stress to gastroenterology trainees in EoE. Greater awareness of burnout and better access to professional support services are needed.

## Data Availability

Supplementary data available upon reasonable request.

## Acknowledgements

This research was not funded however communications and access to literature were provided by the University of Cambridge. JO is funded by the W.D. Armstrong Doctoral Research Training Fellowship and a talent development grant from the National University of Singapore.

## Author’s contributions

Conceptualisation: JO, design: JO and CS, data curation JO and CS, statistical analysis: JO, manuscript preparation: JO, CS, LWY, revisions: SO, YAN, critical edits and supervision: AS.

## Data sharing

No additional data is available

---

**Supplementary Table 1:** Burnout Questionnaire Part 2 - Free text questions.

| *Please fill in the blanks* |  |  |
| --- | --- | --- |
| S/N | Question | Response |
| 1 | Age: |  |
| 2 | Gender: |  |
| 3 | Grade (e.g. ST3, clinical research fellow etc.): |  |
| 4 | Are you in fulltime or less than full time training?: |  |
| 5 | Do you work in a district hospital, tertiary centre or research centre?: |  |
| 6 | Do you do General Internal Medicine oncalls?: |  |
| 7 | Which area of your life contributes most to your stress levels (e.g. job, personal life, finances etc)?: |  |
| 8 | Which job-related factor contributes most to your stress levels (e.g. exams, workload etc)?: |  |
| 9 | Are you aware of any professional support services for trainees in the deanery?: |  |

## Go straight to content

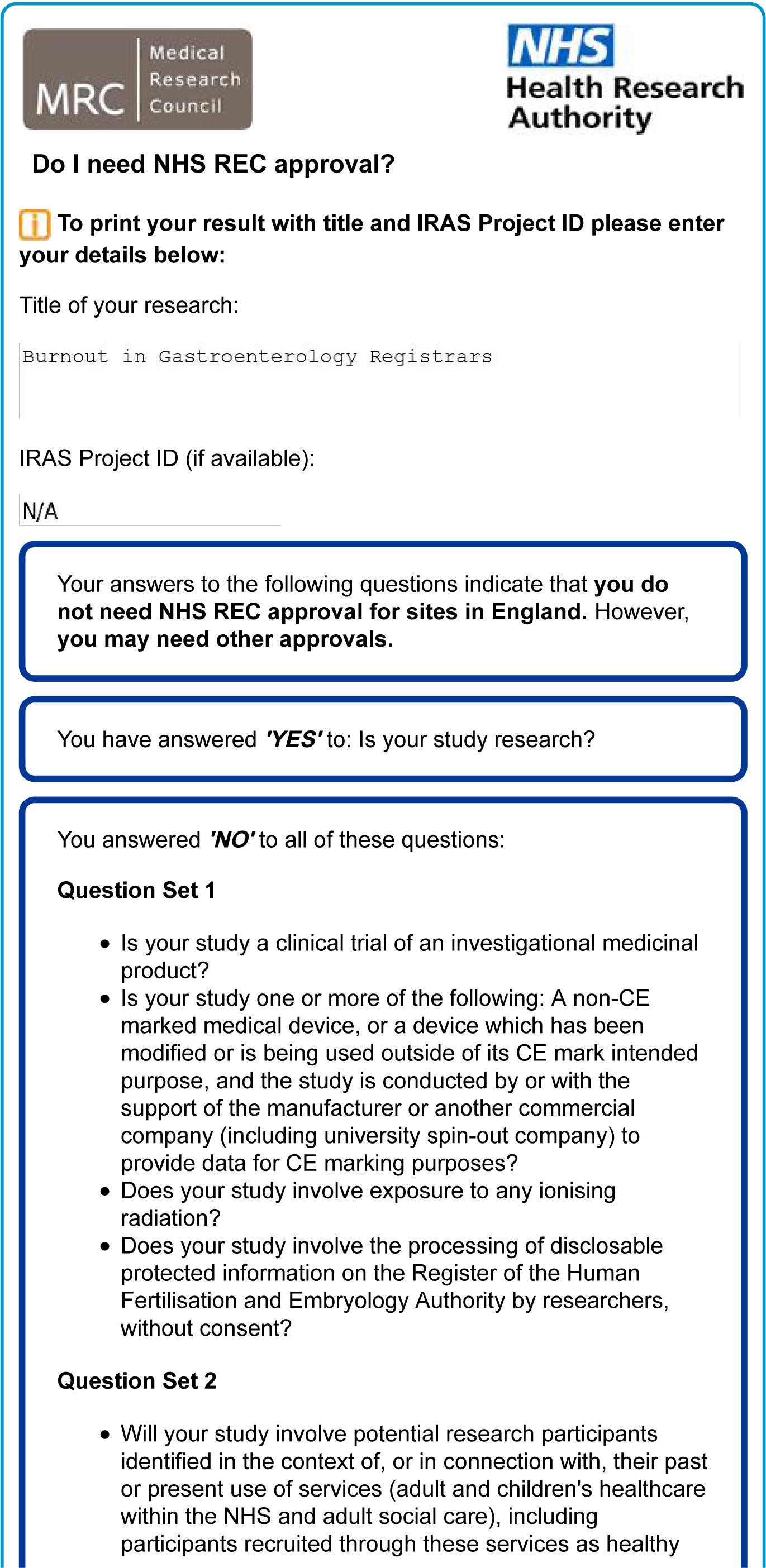
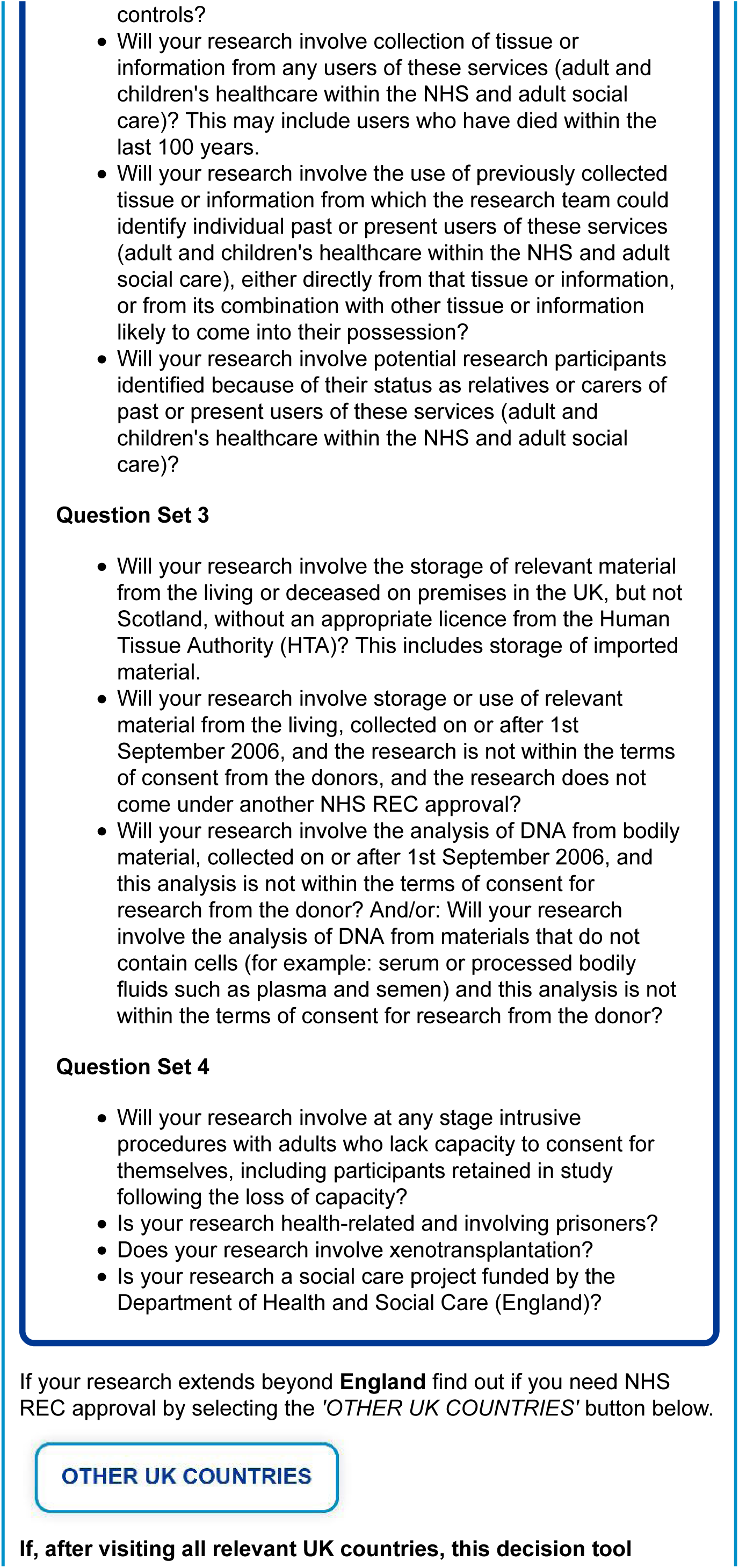
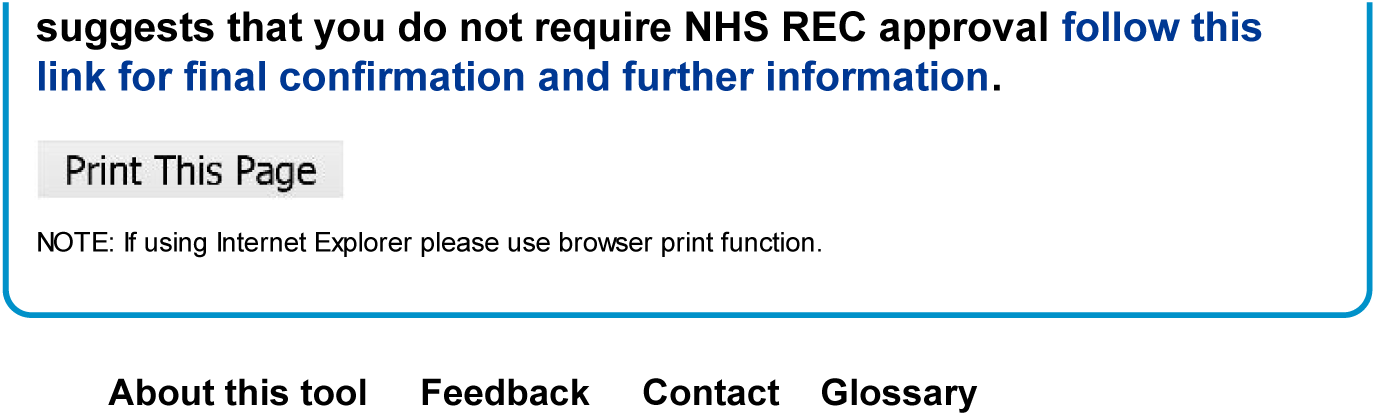

## STROBE 2007 (v4) Statement—Checklist of items that should be included in reports of *cross-sectional studies*

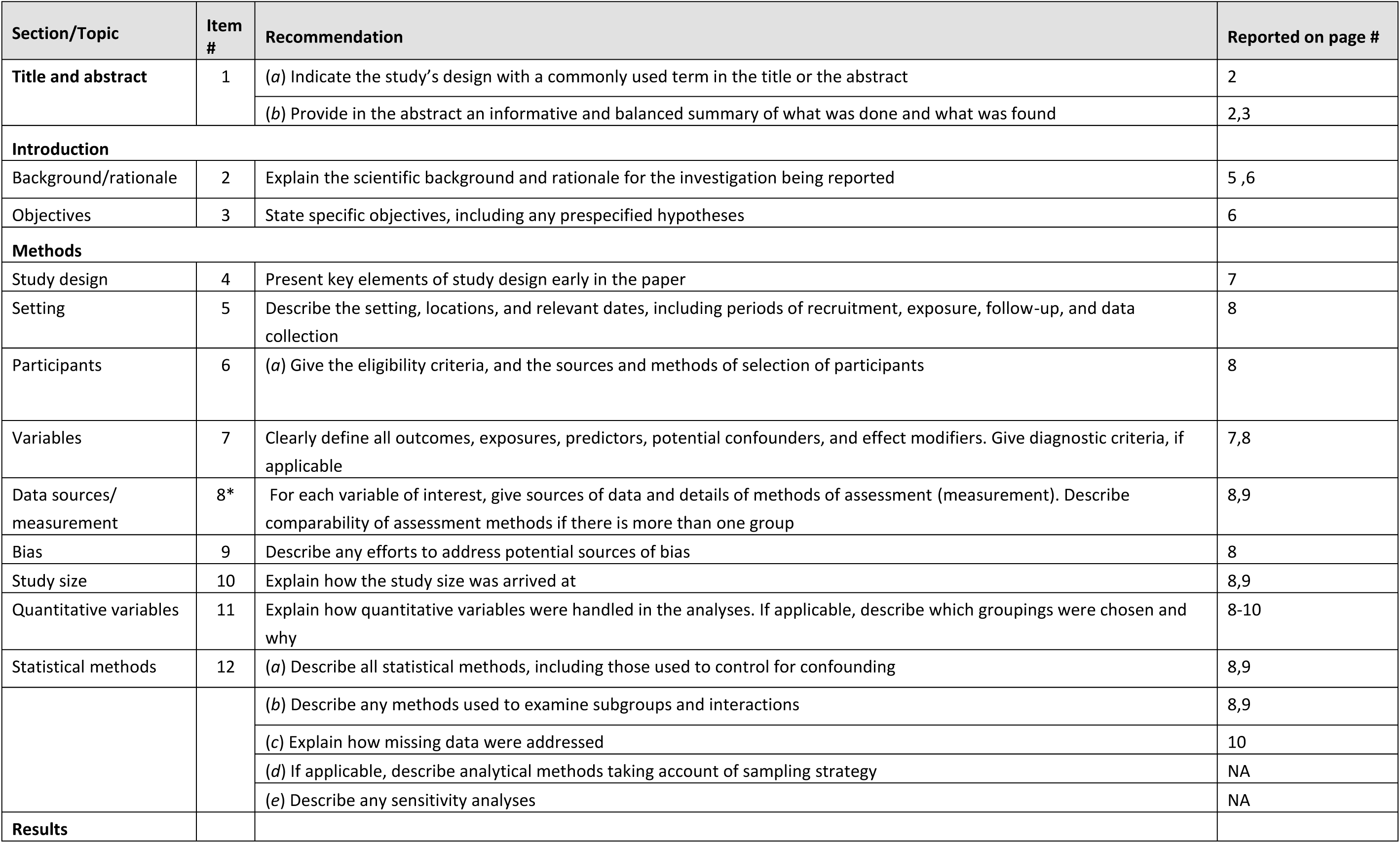
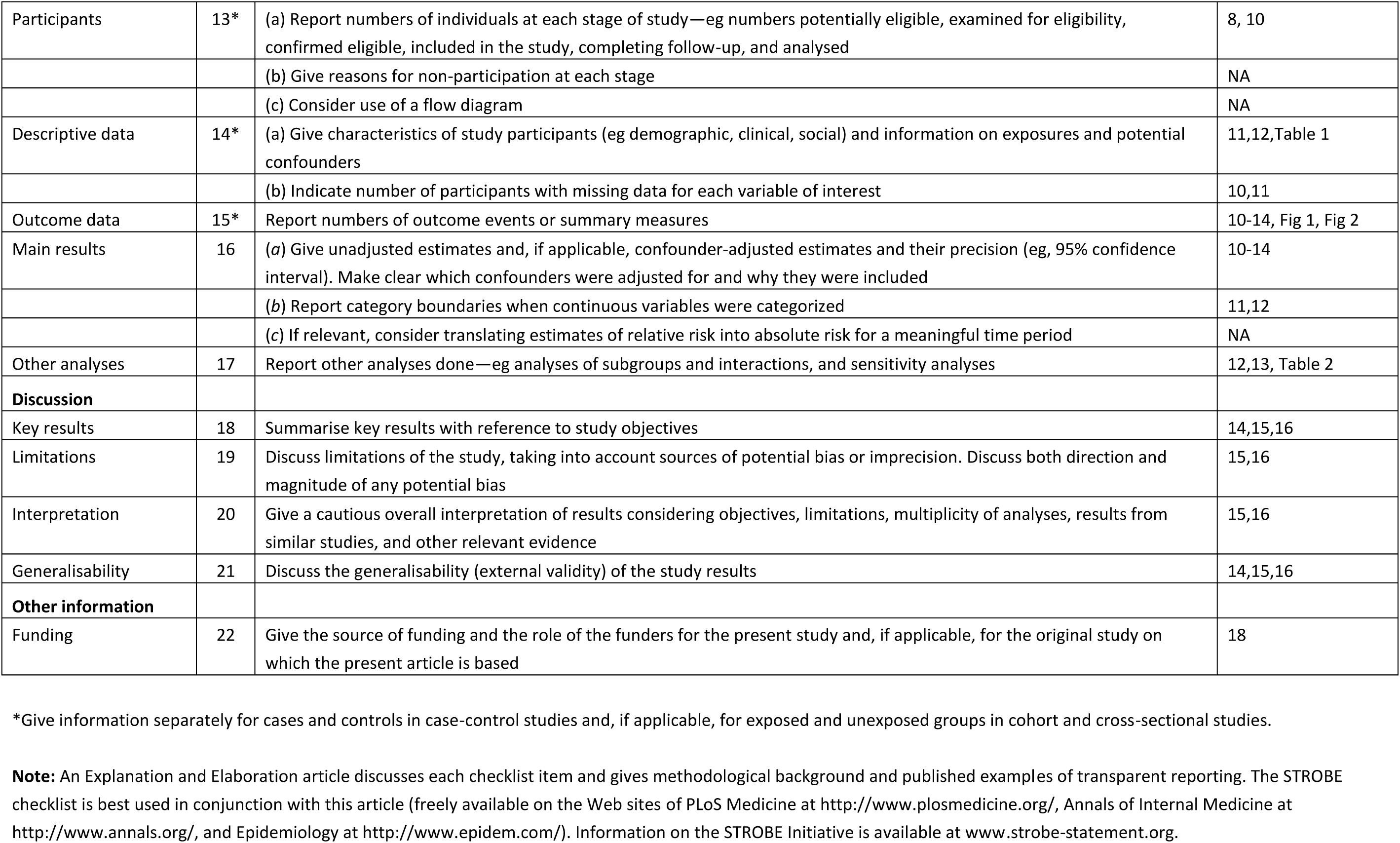

